# Prospective Analysis of urINe LAM to Eliminate NTM Sputum Screening (PAINLESS) study: Rationale and trial design for testing urine lipoarabinomannan as a marker of NTM lung infection in cystic fibrosis

**DOI:** 10.1101/2024.08.08.24311698

**Authors:** Kara M. Calhoun, Emily Armantrout, Katie Poch, Silvia Caceres, Valerie K. Lovell, Marion Jones, Kenneth C. Malcolm, Brian Vestal, Emily Wheeler, Noel Rysavy, Jordan Manzer, Ibrahim Aboellail, Delphi Chatterjee, Jerry A. Nick

## Abstract

**Background:** Routine screening for nontuberculous mycobacterial (NTM) lung disease is dependent on sputum cultures. This is particularly challenging in the cystic fibrosis (CF) population due to reduced sputum production and low culture sensitivity. Biomarkers of infection that do not rely on sputum may lead to earlier diagnosis, but validation trials require a unique prospective design.

**Purpose:** The rationale of this trial is to investigate the utility of urine lipoarabinomannan (LAM) as a test to identify people with CF with a new positive NTM culture. We hypothesize that urine LAM is a sensitive, non-invasive screening test with a high negative predictive value to identify individuals with a relatively low risk of having positive NTM sputum culture.

**Study design:** This is a prospective, single-center, non-randomized observational study in adults with CF, 3 years of negative NTM cultures, and no known history of NTM positive cultures. Patients are followed for two year-long observational periods with the primary endpoint being a positive NTM sputum culture within a year of a positive urine LAM result and a secondary endpoint of a positive NTM sputum culture within 3 years of a positive urine LAM result. Study implementation includes remote consent and sample collection to accommodate changes from the COVID-19 pandemic.

**Conclusions:** This report describes the study design of an observational study aimed at using a urine biomarker to assist in the diagnosis of NTM lung infection in pwCF. If successful, urine LAM could be used as an adjunct to traditional sputum cultures for routine NTM screening.

## INTRODUCTION

People with cystic fibrosis (pwCF) are a population at risk for developing pulmonary nontuberculous mycobacteria (NTM) infection, and despite recent advances in the field, diagnosis and treatment remains challenging (1-3). Historically, approximately 20% of cystic fibrosis (CF) patients have a positive NTM culture over a 5 year period (4); thus, annual screening for NTM is recommended (3). *M. avium* complex (MAC) and *M. abscessus* (MABSC) are responsible for nearly all NTM infections in pwCF living in the US (5). Both species can cause NTM lung disease, which is associated with significant morbidity and mortality.

The diagnosis of NTM lung disease in pwCF is challenging, as co-infections with bacteria such as *P. aeruginosa* and *S. aureus* are common, and may result in identical symptoms and radiographic findings (6). Frequently, NTM may be detected in a culture, but the infection then resolves without treatment or remains indolent for many years. A retrospective review from the Colorado Adult CF Center showed that the majority of participants with a positive NTM culture had transient (23%) or persistent but clinically indolent NTM infection (38.5%) (7). Currently, sputum culture is the “gold standard” for detecting NTM infection and the assay by which all diagnostic and treatment decisions are made (1). However, limitations of sputum culture include slow growth, high cost, and low sensitivity due to the required decontamination steps (3, 8-11). Also, given advancements in CF care and widespread use of cystic fibrosis transmembrane conductance regulator (CFTR) modulator therapy, pwCF are increasingly unable to routinely expectorate sputum (12) and are presenting less often to clinic or with pulmonary exacerbations (13). This combination of factors highlights the need for sputum-independent markers of NTM infection in this population (14).

Lipoarabinomannan (LAM) is a cell wall lipoglycan found in all mycobacteria species and a biomarker of mycobacterial infection (15, 16). During infection, LAM is released from metabolically active or degrading mycobacterial cells into the circulation with subsequent filtration by the kidneys and passed into the urine (17, 18). LAM is a validated biomarker for active tuberculosis (TB) when antigen detection, rather than antibody measurement, is applied (15, 19-23). These antigen detection assays for TB based on LAM are commercially available both as ELISAs and as a point-of-care test. However, the utility of a commercially available TB-specific LAM antigen was tested in a Danish CF clinic and was determined not to be suitable for diagnosis of NTM due to low concentrations of LAM antigen detected in urine from NTM positive pwCF (24). We reported measurable detection of urine LAM utilizing gas chromatography–mass spectrometry (GC-MS) analysis that corresponded with previous NTM culture history in pwCF (n=44) with well-documented NTM status (25). Urine LAM analysis also reflected treatment response in a pwCF receiving bacteriophage therapy for *M. abscessus* (26).

Herein, we present the design of a prospective, single-center, nonrandomized observational study to investigate the utility of urine LAM as a test to identify pwCF with clinically low suspicion for having a sputum culture positive for NTM. The primary objective of this study is to determine the ability of the urine LAM assay to predict continued negative NTM sputum cultures over a 12-month period. Secondary objectives are to determine the ability of the urine LAM assay to predict a new positive NTM sputum culture within 12 months and establish the time between a positive urine LAM assay and a newly positive NTM sputum culture. We hypothesize that urine LAM is a sensitive, non-invasive screening test to identify NTM infection in individuals with a relatively low risk of having a positive NTM sputum culture.

## METHODS

### Study Design

PAINLESS is a prospective, single-center, nonrandomized observational study. The study is approved by the Biomedical Research Alliance of New York (BRANY) review board (20-08-402-528) and is registered in ClinicalTrials.gov (NCT04579211). We comply with the Declaration of Helsinki and Good Clinical Practice guidelines. Written or electronic informed consent is obtained from all participants with a target enrollment of 100. As subject enrollment began on October 23 2020 and continued during restricted clinic access due to the Covid-19 pandemic, informed consent remains available by utilizing an innovative, electronic consenting platform (ClinOne, Inc). The use of electronic consent eliminates the need to have prolonged in-person contact and decreases the risk of infection from COVID-19 and other viruses/bacteria present in a healthcare setting. An instructional video is available for review prior to participation to further explain the rationale for the study, sample collection procedures, and sample timelines (27). Subjects are provided a urine collection kit to expedite sample drop-off. Subjects who are consented in-person are provided written informed consent and may collect the urine specimen at the time of the visit or at home. Subjects are followed on protocol for a minimum of two year-long observation periods (24±3 months). The trial is currently ongoing. The schedule of events is shown in Figure 1 and an overview of the PAINLESS study design and timeline is shown in Figure 2.

**Figure 1.**
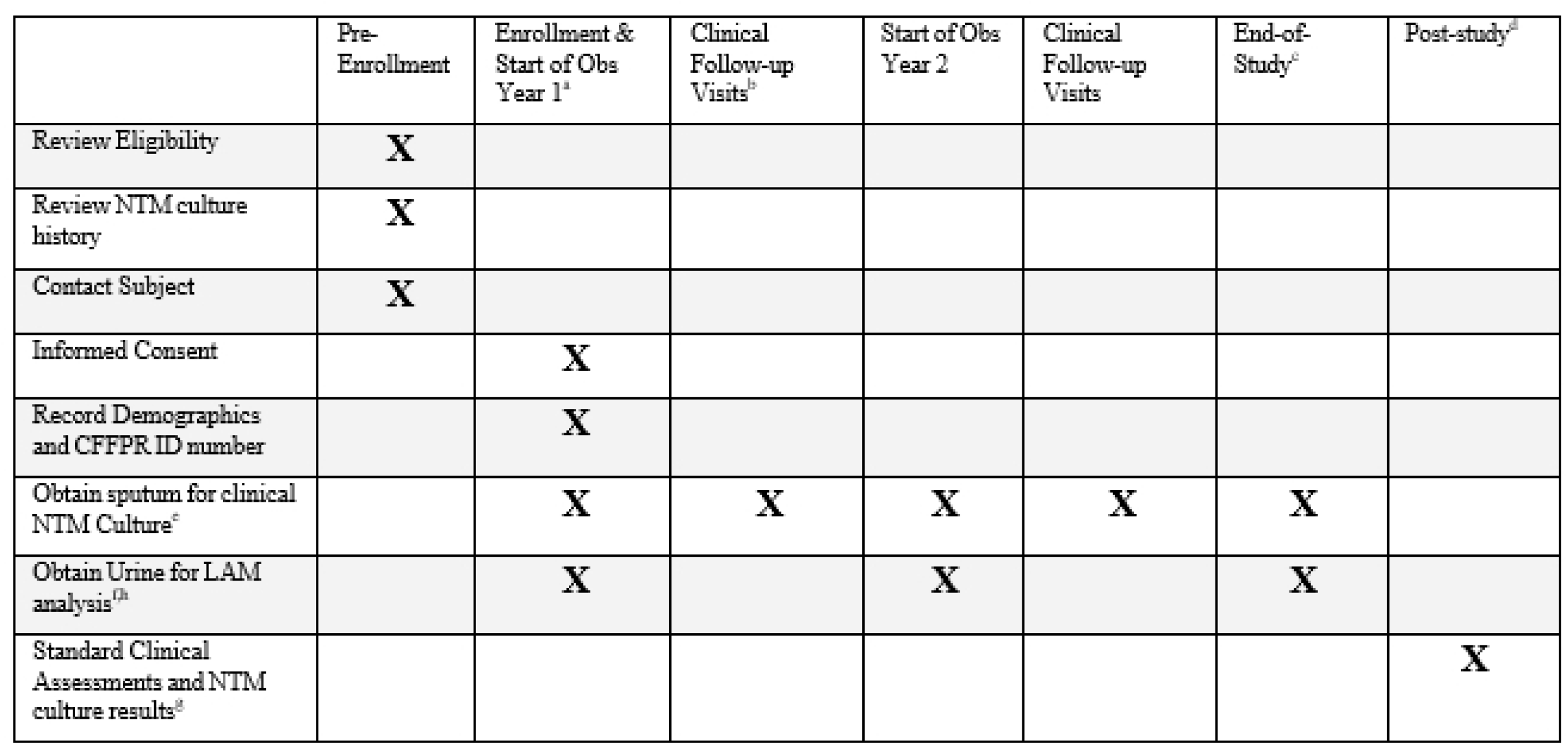
PAINLESS schedule of events. ^a^ Refer to PAINLESS Study Timeline (Figure 2). ^b^ On-study clinical follow-up visits will occur as clinically indicated in clinic, the hospital or at home. ^c^ End-of-study visit will be the first clinical visit occurring > 9 months after the start of Observational Year 2. ^d^ Study personnel will continue to review NTM culture results and clinical events recorded in the CFFPR for up to 5 years following end of study. ^e^ Samples may be obtained in clinic, the hospital or at home, either expectorated or sputum induction. If results from oropharyngeal swab or bronchoscopy are available, they will also be recorded. ^f^ Urine sample must be collected within 30 days of a sputum sample at the start of Year 1 and Year 2. End-of-study urine sample does not need to be linked to a sputum sample. ^g^ Standard clinical assessments will be obtained from CFF patient registry database. ^h^ Replacement urine for LAM assay if needed.

**Figure 2.**
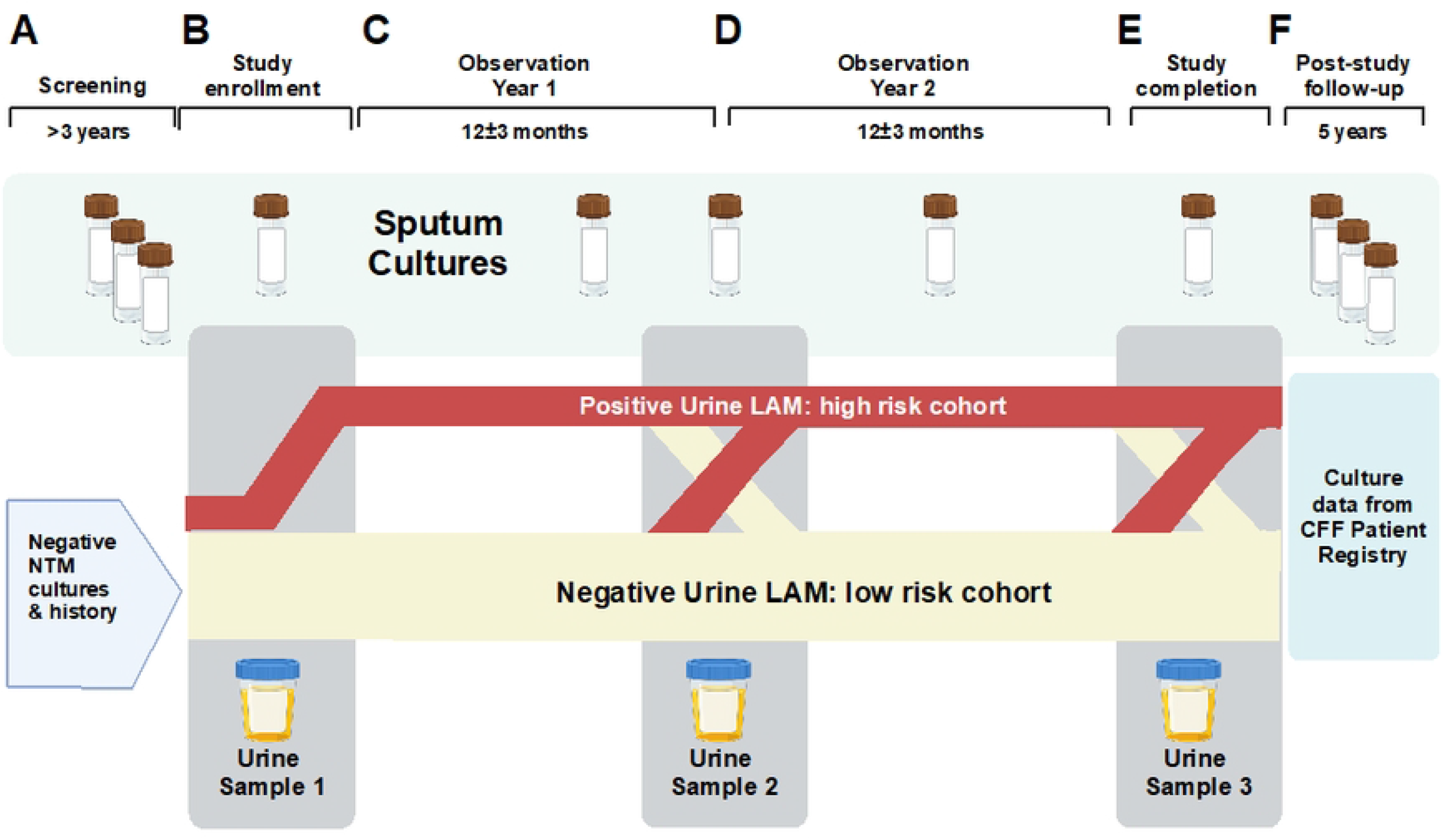
PAINLESS study design and timeline: **Panel A**: Screening criteria requires no history of NTM positive cultures combined with at least 3 negative sputum cultures for NTM over the past 3 years. **Panel B**: At enrollment, the first urine sample for LAM analysis is collected and is within 30 days of a NTM sputum culture. It is anticipated that a positive urine LAM will be detected in a subset of subject, who will comprise a “high risk cohort” that is more likely to have a positive NTM sputum culture. **Panel C**: Over the course of the study, NTM sputum cultures will be obtained when available in the context of clinical care. **Panel D**: At the end of Year 1, a second urine will be collected for LAM analysis. It is anticipated that some subjects with negative LAM at enrollment will convert to positive at each follow-up interval, and likewise some subjects with positive LAM at enrollment will convert to negative. **Panel E**: At the end of the second year of observation a 3^rd^ urine sample will be collected for LAM analysis, as well as sputum for NTM culture (if available). **Panel F**: Results from future sputum NTM cultures will be monitored from the CFF Patient Registry for up to 5 years following the end of study procedures.

### Study population

Adults (≥18 years old) with a confirmed diagnosis of CF followed at the Colorado Adult CF Center who have a minimum of 3 negative NTM cultures with 3 years of available clinical culture data and at least one negative NTM culture within the year prior to enrollment and no known history of previous NTM positive cultures are eligible for enrollment. There are no criteria related to the use of CFTR modulators or other ongoing therapies unrelated to NTM. It is anticipated that up to 90% of participants will be receiving CFTR modulator therapy for the entire duration of the trial, consistent with current usage within the study population. Complete inclusion and exclusion criteria are shown in **Table 1**.

**TABLE 1:** INCLUSION AND EXCLUSION CRITERIA OF THE PAINLESS TRIAL.

| Inclusion Criteria | Exclusion Criteria |
| --- | --- |
| 1. Informed consent obtained online or in person from the participant<br>2. Enrolled in the CFF Patient Registry (CFFPR)<br>3. Be willing and able to adhere to study procedures in the context of clinical care, and other protocol requirements<br>4. Male or female participant ≥ 18 years of age at enrollment who are able expectorate sputum and/or willing to undergo sputum induction (if necessary)<br>5. Diagnosis of CF consistent with the 2017 CFF Guidelines<br>6. NTM culture status of negative, as defined by review of culture data, with at least 3 years of available data; at least 3 NTM negative cultures; and one negative culture within the year prior to enrollment, Also, chart review must confirm no known history of previous positive cultures for pathogenic NTM. | 1. Has any other condition that, in the opinion of the Site Investigator/designee, would preclude informed consent or assent, make study participation unsafe, complicate interpretation of study outcome data, or otherwise interfere with achieving the study objectives |

### Subject related data

Demographic information including date of birth, sex, and race are recorded. The diagnosis of CF consistent with the 2017 CFF Guidelines is confirmed (28). Dates and results from all available NTM cultures are recorded. A minimum of 3 NTM cultures over a 3 year prior to enrollment must be available and documented, with at least 1 negative within the year previous to enrollment. Use of CFTR modulators is also recorded. All subjects in the PAINLESS Trial must also be enrolled in the CF Foundation Patient Registry (CFFPR) (29). If needed, data may be extracted from the CFFPR for exploratory analysis and long-term follow-up.

### Sputum collection and culture analysis

All sputum cultures are collected in the context of clinical care. Expectorated or induced sputum samples are collected during clinic visits and hospitalizations. Sputum samples collected at home may be brought to a clinic appointment or shipped using a sputum collection kit. The sputum collection kits, including instructions for collecting and shipping sputum specimens, are routinely ordered by providers to facilitate sputum culture collection. Samples obtained by bronchoscopy (if available) will also be recorded when collected for clinical indications. Microbiology and mycobiology results are recorded in the subject’s electronic medical record and the CFF Patient Registry. Sputum samples are cultured for NTM as well as typical CF pathogens. NTM cultures are performed at the Advanced Diagnostic Laboratories (ADx) at National Jewish Health, which is a national reference laboratory for mycobacteriology species identification and drug resistance testing (30). Following digestion and decontamination, specimens are inoculated on a LJ Slant, Middlebrook 7H11 Agar/ Mitchison 7H11 Selective Agar biplate(s) and MGIT for up to eight weeks. Suspected NTM growth is confirmed from either liquid and solid culture media using a Ziehl-Neelsen staining. Upon growth detection, genomic DNA is isolated for subsequent PCR amplification. A targeted segment of the DNA-directed RNA polymerase subunit beta (*rpoB*) gene is performed to determine molecular identification.

### Urine collection for LAM analysis

The initial urine sample is obtained at enrollment and marks the start of observational year 1. If the subject is consented remotely, the subject completes the electronic consent process prior to collecting the initial urine sample. The initial urine sample is required within 30 days of a NTM sputum sample collected in the context of clinical care. Following enrollment, additional urine samples are collected at the start of observational year 2 and at the end of study. An additional urine collection may occur in the event a subject is found to have a positive NTM sputum culture between the planned collection dates. The urine sample may be collected in the clinic or at home with a collection kit and brought to clinic. Subjects are asked to provide between 20cc to 50cc of urine per collection. After collection, urine samples centrifuged, aliquoted, and stored at -80ºC until analysis.

### LAM assay methods

All urine samples require a hexane wash (1:1 v/v) to remove exogenous lipids and proteins in order to reduce background and increase the limit of detection. Coded samples are separated by hydrophobic interaction chromatography over Octyl Sepharose (OS)-CL 4B. The 40% & 65% n-propanol in 0.1 M NH_4_OAc eluents off the HIC column are processed for GC/MS analysis as previously described (31, 32). GC/MS analyses are carried out using a Thermo GC-TSQ8000 Evo Triple Quad GC mass spectrometer. Chromatograms with respective peaks are integrated manually (*i*.*e*. peak areas defined manually and integrated areas generated by software) for the estimation of total D-ara and TBSA content. LAM used for standardization and assay validation in this study is purified from *Mtb* and *M. avium* (**Figure 3**) (33).

**Figure 3:**
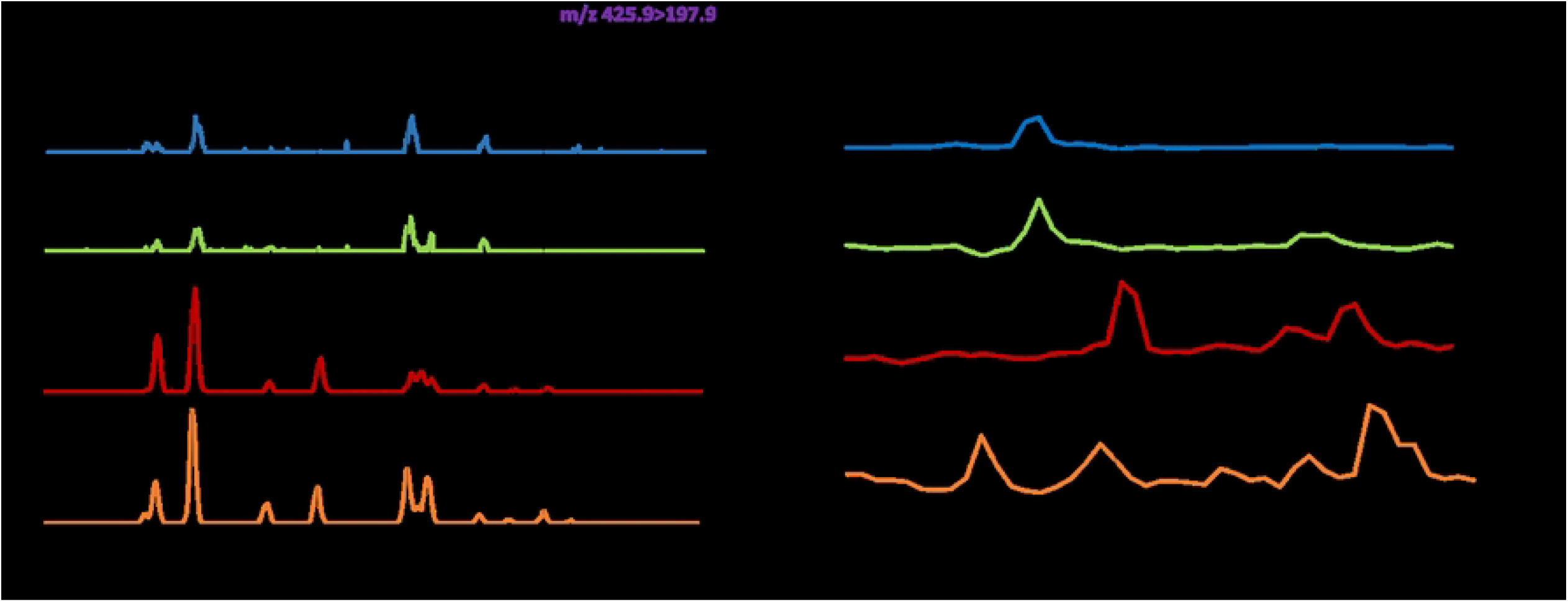
Representative GC/MS chromatograms showing the absence and/or presence of urinary LAM related to NTM-negativity and/or positivity. A) D-Arabinose (D-ara) MS/MS method monitoring m/z 420.9-m/z 192.9): Four characteristic peaks of Internal Standard (^13^C_5_-D-Arabinose; top panel); Sequentially (top to bottom), 309 and 308 NTM negative, D-ara negative; BP201 and 303 are positive, LAM positive, NTM positive. B) Tuberculostearic acid (TBSA) Single ion monitoring (SIM) at m/z 297.3: TBSA standard (C:19; top panel); Sequentially, 309 and 308 NTM negative, TBSA negative; BP201 and 303 are TBSA positive, LAM positive, NTM positive.

## DATA COLLECTION, MANAGEMENT, and ANALYSIS

Study data are collected and managed using the REDCap electronic data capture tools hosted at National Jewish Health. REDCap (Research Electronic Data Capture) is a secure, web-based software platform designed to support data capture for research studies (34, 35). The handling and analysis of the data is conducted using good computing practices for clinical trials, including routine data validation checks. As part of data analysis, the lead biostatistician on the study monitors for data outliers and performs data validation. If data entry errors are encountered, the study database is updated in accordance with the resolved queries. All changes to the study database are documented in an audit trail. Database lock will occur once quality assurance procedures have been completed and will be maintained for 5 years.

### General analysis plan

#### Sample size

Planned sample size is 100 subjects. Based on historical data from the Colorado Adult CF Program, we anticipate approximately 13 subjects will be identified with a new positive NTM culture over the course of two year-long intervals.

#### Primary analysis

The primary analyses will focus on establishing the predictive accuracy of the urine LAM assay compared to both NTM culture results from the day of sampling and aggregated culture results from within a year of the LAM measurement date. As a secondary endpoint, we will focus on how well a positive urine LAM assay predicts a positive NTM culture over a three year period. For the primary endpoints, the classification performance of the LAM assay will be assessed with respect to its ability to predict a positive culture at the time of the test and over the 12-month period prior or following the test. Of primary interest are the test’s sensitivity and specificity, but we will also investigate positive predictive power, negative predictive power, percent-agreement, and Cohen’s Kappa. These values will be computed twice using the binary results of the LAM assay (positive or not-positive) paired with either the binary culture results from the time of the test or with the aggregated results over the study interval (*i*.*e*. ≧1 positive culture versus no positive cultures). We will also investigate if the continuous abundance of LAM is a useful measure by calculating area under the receiver-operator curve, again using culture from the time of the test or aggregated over the 12-month period. Additionally, the multiple repeated observations for each individual in the two 12-month follow-up periods will allow us to perform a time-to-event analysis associating the LAM assay to culture results. Here, Cox proportional hazards regression models will be utilized with time to first positive NTM culture as the outcome and LAM assay (using both the continuous and binary versions) as the time-varying covariate of interest.

## DISCUSSION

Consensus recommendations call for annual NTM screening for pwCF who can produce sputum, as well as screening before and six months after beginning chronic azithromycin therapy and annually thereafter (3). Screening for NTM infection in the CF population is currently reliant on traditional sputum culture techniques, as oropharyngeal swabs are not recommended due to low sensitivity. Sputum sampling has long been challenging in pwCF who do not produce sputum, and reliance on sputum cultures has become even less feasible with widespread adoption of CFTR modulator therapy. The CFTR triple combination therapy of elexacaftor, tezacaftor, and ivacaftor (E/T/I) was approved in 2019 in the U.S. for pwCF who are either homozygous (36) or heterozygous for F508del mutation (37). A reduction in sputum production is a nearly universal benefit of E/T/I (12). The 2022 CFFPR reported an overall decrease in both microbiology and mycobacterial cultures reported for the last three years These decreases can be attributed to not only the benefits of CFTR modulator therapy, but reduced frequency of in-person care as a result of the COVID-19 pandemic (35). However, the ability of a pwCF to produce a sputum sample has remained steady at 70% since the introduction of E/T/I, though it begins to drop after the age of 30 years. As the life expectancy of pwCF continues to rise, it can be expected that fewer and fewer pwCF will be able to spontaneously expectorate sputum and yields from induced sputum will fall as well. Thus, it is widely recognized that sputum-independent, non-culture-based markers for NTM are an unmet need in CF care. (14, 38).

Airway infection with NTM are among the most serious and least understood comorbidities of CF lung disease, with some patients developing rapid progression of disease. These patients may potentially benefit from an early eradication approach to treatment. PAINLESS is a single-center pilot trial created to investigate the utility of urine LAM as a screening test for the majority of pwCF who will likely never have a sputum culture positive for NTM. Results from this study will determine the feasibility of a larger multi-center trial to test this noninvasive urine assay as a strategy to eliminate the need for routine sputum screening for NTM. A positive urine LAM would notify clinicians of the need for airway cultures and close follow-up, while a negative result would assure the patient, family, and care team that in the absence of clinical suspicion, additional costly and invasive testing is not needed. Given the relatively low sensitivity of NTM cultures, the potential exists that when positive, the urine LAM assay would result in earlier detection than annual sputum screening and serve as an important component to an evidence-based eradication protocol, which currently does not exist. This study also serves as a platform for the development of a more accessible ELISA-based assay for the detection of urine LAM to be potentially used widely in clinical practice. This trial is the first to our knowledge for noninvasive screening of NTM infection in the CF population and also pilots aspects of remote consent and sample collection.

The PAINLESS trial has several limitations that are inherent to the identification and validation of biomarkers. Assessment of urine LAM as an assay is linked to sputum cultures, which are insensitive in this setting, and will often not be available. Thus, in some cases positive urine LAM values will be incorrectly assessed as false positives, when in fact they may be a more sensitive marker than the “gold standard” sputum culture. In addition, it is well recognized that NTM may be transient in the CF population (7), and this phenomena may also result in an apparent false positive urine LAM result. Finally, the current trial utilizes GC/MS analysis of urine LAM, which is not yet standardized. Given these limitation, the greatest value of urine LAM may be to identify those who don’t have an NTM infection. Urine LAM collection for the current trial will end in 2024, with a follow-up period needed to assess sputum culture results. Full validation of the method will require a larger, prospective trial including both children and adults over an extended duration.

## Data Availability

No datasets were generated or analysed during the current study. All relevant data from this study will be made available upon study completion.

NA

## Abbreviations

CF: cystic fibrosis
NTM: nontuberculous mycobacteria
pwCF: people with cystic fibrosis
CFTR: cystic fibrosis transmembrane conductance regulator
LAM: lipoarabinomannan
MAC: *Mycobacterium avium* complex
MABSC: *Mycobacterium abscessus*

## Works Cited

1. Griffith DE, Aksamit T, Brown-Elliott BA, Catanzaro A, Daley C, Gordin F, Holland SM, Horsburgh R, Huitt G, Iademarco MF, Iseman M, Olivier K, Ruoss S, von Reyn CF, Wallace RJ, Jr., Winthrop K. An official ATS/IDSA statement: diagnosis, treatment, and prevention of nontuberculous mycobacterial diseases. Am J Respir Crit Care Med 2007; 175: 367–416.

2. Daley CL, Iaccarino JM, Lange C, Cambau E, Wallace RJ, Andrejak C, Bottger EC, Brozek J, Griffith DE, Guglielmetti L, Huitt GA, Knight SL, Leitman P, Marras TK, Olivier KN, Santin M, Stout JE, Tortoli E, van Ingen J, Wagner D, Winthrop KL. Treatment of Nontuberculous Mycobacterial Pulmonary Disease: An Official ATS/ERS/ESCMID/IDSA Clinical Practice Guideline. Clin Infect Dis 2020; 71: 905–913.

3. Floto RA, Olivier KN, Saiman L, Daley CL, Herrmann JL, Nick JA, Noone PG, Bilton D, Corris P, Gibson RL, Hempstead SE, Koetz K, Sabadosa KA, Sermet-Gaudelus I, Smyth AR, van Ingen J, Wallace RJ, Winthrop KL, Marshall BC, Haworth CS. US Cystic Fibrosis Foundation and European Cystic Fibrosis Society consensus recommendations for the management of non-tuberculous mycobacteria in individuals with cystic fibrosis. Thorax 2016; 71 Suppl 1: i1–i22.

4. Adjemian J, Olivier KN, Prevots DR. Epidemiology of Pulmonary Nontuberculous Mycobacterial Sputum Positivity in Patients with Cystic Fibrosis in the United States, 2010-2014. Annals of the American Thoracic Society 2018; 15: 817–826.

5. Adjemian J, Olivier KN, Prevots DR. Nontuberculous mycobacteria among patients with cystic fibrosis in the United States: screening practices and environmental risk. Am J Respir Crit Care Med 2014; 190: 581–586.

6. Martiniano S, Ja N, Daley C. Nontuberculous Mycobacterial Infections in Cystic Fibrosis. Clin Chest Med 2022; 43: 697–716.

7. Martiniano SL, Sontag MK, Daley CL, Nick JA, Sagel SD. Clinical significance of a first positive nontuberculous mycobacteria culture in cystic fibrosis. Annals of the American Thoracic Society 2014; 11: 36–44.

8. Buijtels PC, Petit PL. Comparison of NaOH-N-acetyl cysteine and sulfuric acid decontamination methods for recovery of mycobacteria from clinical specimens. Journal of microbiological methods 2005; 62: 83–88.

9. Brown-Elliott BA, Griffith DE, Wallace RJ, Jr. Diagnosis of nontuberculous mycobacterial infections. Clinics in laboratory medicine 2002; 22: 911–925, vi.

10. Steingart KR, Ng V, Henry M, Hopewell PC, Ramsay A, Cunningham J, Urbanczik R, Perkins MD, Aziz MA, Pai M. Sputum processing methods to improve the sensitivity of smear microscopy for tuberculosis: a systematic review. The Lancet Infectious diseases 2006; 6: 664–674.

11. Bange FC, Bottger EC. Improved decontamination method for recovering mycobacteria from patients with cystic fibrosis. European journal of clinical microbiology & infectious diseases : official publication of the European Society of Clinical Microbiology 2002; 21: 546–548.

12. Nichols DP, Morgan SJ, Skalland M, Vo AT, Van Dalfsen JM, Singh SB, Ni W, Hoffman LR, McGeer K, Heltshe SL, Clancy JP, Rowe SM, Jorth P, Singh PK, Group PR-MS. Pharmacologic improvement of CFTR function rapidly decreases sputum pathogen density, but lung infections generally persist. J Clin Invest 2023; 133.

13. Marshall B. Cystic Fibrosis Foundation Patient Registry: 2022 Annual Data Report. Bethesda, Maryland ©2023 Cystic Fibrosis Foundation 2023.

14. Nick JA, Malcolm KC, Hisert KB, Wheeler EA, Rysavy NM, Poch K, Caceres S, Lovell VK, Armantrout E, Saavedra MT, Calhoun K, Chatterjee D, Aboellail I, De P, Martiniano SL, Jia F, Davidson RM. Culture independent markers of nontuberculous mycobacterial (NTM) lung infection and disease in the cystic fibrosis airway. Tuberculosis 2023; 138: 102276.

15. Amin AG, D. P, Spencer JS, Brennan PJ, Daum J, Andre BG, Joe M, Bai Y, Laurentius L, Porter MD, Honnen WJ, Choudhary A, Lowary TL, Pinter A, Chatterjee D. Detection of lipoarabinomannan in urine and serum of HIV-positive and HIV-negative TB suspects using an improved capture-enzyme linked immuno absorbent assay and gas chromatography/mass spectrometry. Tuberculosis 2018; 111: 178–187.

16. Torrelles JB, Chatterjee D. Collected Thoughts on Mycobacterial Lipoarabinomannan, a Cell Envelope Lipoglycan. Pathogens 2023; 12.

17. Minion J, Leung E, Talbot E, Dheda K, Pai M, Menzies D. Diagnosing tuberculosis with urine lipoarabinomannan: systematic review and meta-analysis. Eur Respir J 2011; 38: 1398–1405.

18. Lawn SD. Point-of-care detection of lipoarabinomannan (LAM) in urine for diagnosis of HIV-associated tuberculosis: a state of the art review. BMC Infect Dis 2012; 12: 103.

19. Gupta-Wright A, Peters JA, Flach C, Lawn SD. Detection of lipoarabinomannan (LAM) in urine is an independent predictor of mortality risk in patients receiving treatment for HIV-associated tuberculosis in sub-Saharan Africa: a systematic review and meta-analysis. BMC medicine 2016; 14: 53.

20. Lawn SD, Kerkhoff AD, Nicol MP, Meintjes G. Underestimation of the True Specificity of the Urine Lipoarabinomannan Point-of-Care Diagnostic Assay for HIV-Associated Tuberculosis. Journal of acquired immune deficiency syndromes 2015; 69: e144–146.

21. Sigal GB, Pinter A, Lowary TL, Kawasaki M, Li A, Mathew A, Tsionsky M, Zheng RB, Plisova T, Shen K, Katsuragi K, Choudhary A, Honnen WJ, Nahid P, Denkinger CM, Broger T. A Novel Sensitive Immunoassay Targeting the 5-Methylthio-d-Xylofuranose-Lipoarabinomannan Epitope Meets the WHO’s Performance Target for Tuberculosis Diagnosis. J Clin Microbiol 2018; 56.

22. Broger T, Tsionksy M, Mathew A, Lowary TL, Pinter A, Plisova T, Bartlett D, Barbero S, Denkinger CM, Moreau E, Katsuragi K, Kawasaki M, Nahid P, Sigal GB. Sensitive electrochemiluminescence (ECL) immunoassays for detecting lipoarabinomannan (LAM) and ESAT-6 in urine and serum from tuberculosis patients. PLoS One 2019; 14: e0215443.

23. MacLean E, Broger T, Yerlikaya S, Fernandez-Carballo BL, Pai M, Denkinger CM. A systematic review of biomarkers to detect active tuberculosis. Nature microbiology 2019; 4: 748–758.

24. Qvist T, Johansen IS, Pressler T, Hoiby N, Andersen AB, Katzenstein TL, Bjerrum S. Urine lipoarabinomannan point-of-care testing in patients affected by pulmonary nontuberculous mycobacteria--experiences from the Danish Cystic Fibrosis cohort study. BMC Infect Dis 2014; 14: 655.

25. De P, Amin AG, Graham B, Martiniano SL, Caceres SM, Poch KR, Jones MC, Saavedra MT, Malcolm KC, Nick JA, Chatterjee D. Urine lipoarabinomannan as a marker for low-risk of NTM infection in the CF airway. Journal of cystic fibrosis : official journal of the European Cystic Fibrosis Society 2020; 19: 801–807.

26. Nick JA, Dedrick RM, Gray AL, Vladar EK, Smith BE, Freeman KG, Malcolm KC, Epperson LE, Hasan NA, Hendrix J, Callahan K, Walton K, Vestal B, Wheeler E, Rysavy NM, Poch K, Caceres S, Lovell VK, Hisert KB, de Moura VC, Chatterjee D, De P, Weakly N, Martiniano SL, Lynch DA, Daley CL, Strong M, Jia F, Hatfull GF, Davidson RM. Host and pathogen response to bacteriophage engineered against Mycobacterium abscessus lung infection. Cell 2022; 185: 1860–1874 e1812.

27. Lovell V, Caceres S. Introduction to the Prospective Analysis of urINe LAM to Eliminate NTM Sputum Screening (PAINLESS) study. 2020. Available from: https://www.dropbox.com/s/evaeivjey28mfab/V3.mp4?dl=0.

28. Farrell PM, White TB, Ren CL, Hempstead SE, Accurso F, Derichs N, Howenstine M, McColley SA, Rock M, Rosenfeld M, Sermet-Gaudelus I, Southern KW, Marshall BC, Sosnay PR. Diagnosis of Cystic Fibrosis: Consensus Guidelines from the Cystic Fibrosis Foundation. J Pediatr 2017; 181S: S4–S15 e11.

29. Knapp EA, Fink AK, Goss CH, Sewall A, Ostrenga J, Dowd C, Elbert A, Petren KM, Marshall BC. The Cystic Fibrosis Foundation Patient Registry. Design and Methods of a National Observational Disease Registry. Annals of the American Thoracic Society 2016; 13: 1173–1179.

30. Khare R. Advanced Diagnostic Laboratories Mycobacteriology Laboratory. 2024 [cited 2024 May 26]. Available from: https://www.nationaljewish.org/for-professionals/diagnostic-testing/advanced-diagnostic-laboratories/our-laboratories/mycobacteriology.

31. De P, Amin AG, Valli E, Perkins MD, McNeil M, Chatterjee D. Estimation of D-Arabinose by Gas Chromatography/Mass Spectrometry as Surrogate for Mycobacterial Lipoarabinomannan in Human Urine. PLoS One 2015; 10: e0144088.

32. Manzer J, Aboellail I, Ja N, Chatterjee D, Amin AG. Urine Lipoarabinomannan as a Biomarker for Mycobacterium tuberculosis and non-tuberculous mycobacterial infections. protocolsio 2024; May 31, 2024.

33. Shi L, Torrelles JB, Chatterjee D. Lipoglycans of Mycobacterium tuberculosis: isolation, purification, and characterization. In: Brown TPaAC, editor. Mycobacteria Protocols, second edition ed. Barts and The London, London, UK: Humana Press; 2009. p. 23–45.

34. Harris PA, Taylor R, Minor BL, Elliott V, Fernandez M, O’Neal L, McLeod L, Delacqua G, Delacqua F, Kirby J, Duda SN, Consortium RE. The REDCap consortium: Building an international community of software platform partners. J Biomed Inform 2019; 95: 103208.

35. Harris PA, Taylor R, Thielke R, Payne J, Gonzalez N, Conde JG. Research electronic data capture (REDCap)--a metadata-driven methodology and workflow process for providing translational research informatics support. J Biomed Inform 2009; 42: 377–381.

36. Heijerman HGM, McKone EF, Downey DG, Van Braeckel E, Rowe SM, Tullis E, Mall MA, Welter JJ, Ramsey BW, McKee CM, Marigowda G, Moskowitz SM, Waltz D, Sosnay PR, Simard C, Ahluwalia N, Xuan F, Zhang Y, Taylor-Cousar JL, McCoy KS, Group VXT. Efficacy and safety of the elexacaftor plus tezacaftor plus ivacaftor combination regimen in people with cystic fibrosis homozygous for the F508del mutation: a double-blind, randomised, phase 3 trial. Lancet 2019; 394: 1940–1948.

37. Middleton PG, Mall MA, Drevinek P, Lands LC, McKone EF, Polineni D, Ramsey BW, Taylor-Cousar JL, Tullis E, Vermeulen F, Marigowda G, McKee CM, Moskowitz SM, Nair N, Savage J, Simard C, Tian S, Waltz D, Xuan F, Rowe SM, Jain R, Group VXS. Elexacaftor-Tezacaftor-Ivacaftor for Cystic Fibrosis with a Single Phe508del Allele. The New England journal of medicine 2019; 381: 1809–1819.

38. van Ingen J, Aksamit T, Andrejak C, Bottger EC, Cambau E, Daley CL, Griffith DE, Guglielmetti L, Holland SM, Huitt GA, Koh WJ, Lange C, Leitman P, Marras TK, Morimoto K, Olivier KN, Santin M, Stout JE, Thomson R, Tortoli E, Wallace RJ, Jr., Winthrop KL, Wagner D, for N-N. Treatment outcome definitions in nontuberculous mycobacterial pulmonary disease: an NTM-NET consensus statement. Eur Respir J 2018; 51.

